# Mediterranean-DASH Intervention for Neurodegenerative Delay diet and risk of dementia: three prospective studies and meta-analysis of cohort studies

**DOI:** 10.1101/2022.10.16.22281155

**Authors:** Hui Chen, Klodian Dhana, Yuhui Huang, Liyan Huang, Yang Tao, Xiaoran Liu, Debora Melo van Lent, Yan Zheng, Alberto Ascherio, Walter Willett, Changzheng Yuan

**Affiliations:** School of Public Health and the Second Affiliated Hospital, Zhejiang University School of Medicine, Hangzhou, China; Rush Institute for Healthy Aging, Rush University Medical Center, Chicago, Illinois, USA; Department of Internal Medicine, Rush University Medical Center, Chicago, Illinois, USA; Glenn Biggs Institute for Alzheimer’s & Neurodegenerative Diseases, UT Health San Antonio, San Antonio, TX, USA; Department of Neurology, Boston University School of Medicine, Boston, MA, USA; Framingham Heart Study, Framingham, MA, USA; State Key Laboratory of Genetic Engineering, Human Phenome Institute, and School of Life Sciences, Fudan University, Shanghai, China; Department of Nutrition, Harvard T H Chan School of Public Health, Boston, Massachusetts, USA; Channing Division of Network Medicine, Brigham and Women’s Hospital and Harvard Medical School, Boston, Massachusetts, USA

## Abstract

**Objective:** To evaluate the association of Mediterranean-DASH Intervention for Neurodegenerative Delay (MIND) diet with risk of dementia in three prospective cohort studies, and to conduct a meta-analysis of cohort studies.

**Design & Setting:** Three prospective cohort studies – the Whitehall II study (WII, mean follow-up=13.2 years), the Health and Retirement Study (HRS, mean follow-up=4.3 years) and Framingham Heart Study Offspring cohort (FOS, mean follow-up=10.4 years), and meta-analysis of cohort studies.

**Participants:** We included 18163 middle-aged and older women and men (8360 in WII, 6758 in HRS, and 3045 in FOS) without baseline prevalent dementia for cohort analyses.

**Main outcome measures:** Incident all-cause dementia, with cohort-specific definitions.

**Results:** The mean (SD) of baseline MIND diet score (range: 0-15) was 8.3 (1.4) in WII, 7.1 (1.9) in HRS, and 8.1 (1.6) in FOS. Over 171,079 person-years, a total of 802 participants (222 in WII, 338 in HRS, and 242 in FOS) developed incident dementia. In the multivariable-adjusted model, every 3-point increment in MIND diet score (range: 0-15) was related to 20% decreased risk of dementia (pooled hazard ratio (HR) = 0.80; 95% confidence interval (CI): 0.70-0.91, P-trend=0.0014, P-heterogeneity=0.3039). In the WII, the corresponding HRs (95% CIs) was 1.04 (0.71-1.52) for remote MIND score in 1991-1993, and 0.76 (0.53-1.09) for recent score in 2002-2004. In the FOS, the HRs were 0.66 (0.50-0.86) for MIND score in 1991-1995 and 0.67 (0.51-0.88) for that in 1998-2001, respectively. In the meta-analysis of eleven cohorts with 224,140 participants, the highest category of MIND diet score was related to 18% lower risk of dementia (pooled relative risk: 0.82, 95% CI: 0.74-0.92, I^2^=74%, P-heterogeneity<0.01) compared with the lowest category.

**Conclusions:** Adherence to the MIND diet was related to lower risk of incident dementia in middle-aged and older adults. Further studies are warranted to develop and refine the specific MIND diet for different populations.

**Systematic review registration:** INPLASY202270127

**Summary box:** *What is already known on this topic:*

- Mediterranean-DASH Intervention for Neurodegenerative Delay (MIND) diet has been proposed as a healthy dietary pattern for human brain.
- The relation of MIND to incident dementia was relatively inconsistent and inconclusive in previous cohort studies.

*What this study adds:*

- Results from the cohort analyses and the meta-analysis confirmed that adherence to the MIND diet is prospectively related to lower risk of incident dementia.
- Results in current studies are heterogeneous across different populations, calling for further investigations into the culture-specific MIND diet.

## Introduction

All-cause dementia increasingly burdens healthcare systems and threatens the well-being of older adults [1,2], and lack of effect treatments makes its early prevention of great importance[1,3]. Among numerous potential risk factors[1,4], dietary factors have aroused major interest[5–8], including specific nutrients[9–13], food groups[14,15], and dietary patterns [6,8,16].

Devised by Morris et al.[17], Mediterranean-DASH Intervention for Neurodegenerative Delay (MIND) diet was associated with lower risk of Alzheimer’s disease (AD) [18] and slower cognitive decline[17]. The MIND diet emphasizes natural plant-based foods and limited intakes of animal and high saturated fat foods and uniquely encourages consumption of berries and green leafy vegetables[17]. Although studies have assessed its relation to cognitive function and decline [17,19,20], only several studies have linked it to all-cause dementia or Alzheimer’s Disease (AD) dementia, with inconclusive results. A prospective study in 923 US participants aged 58 to 98 years (mean follow-up=4.5 years) found that higher adherence to MIND diet was related to lower risk of AD dementia [18]. In the Rotterdam Study [21], the relation of the MIND diet to incident all-cause was significant in the first 7 years of follow-up, but was non-significant afterwards. In aggregate, despite its biological plausibility [22], population-based evidence for the role of the MIND diet in dementia prevention is inconclusive.

In the current study, we assessed the associations of adherence to the MIND diet with incident all-cause dementia in the Whitehall II study (WII) in UK and the Health and Retirement Study (HRS) and the Framingham Heart Study Offspring cohort (FOS) in the US. Further, we conducted a meta-analysis summing all current evidence on the relation of MIND diet to dementia.

## Methods

### 1. Cohort analyses

#### Study population

We leveraged individual-level data from three prospective studies. The WII was aimed at understanding the causes of age-related heterogeneity in health[23]. WII enrolled 10,308 civil working women and men aged 35-55 at baseline (1985), followed by eleven data collection phases to date. Participants finished repeated dietary assessments [24] at Phase 3 (1991-1993), 5 (1997-1999), and 7 (2002-2004). The HRS is a nationally representative cohort study of 30,000 adults aged 50 and older in the USA[25,26]. In 2013, 8,035 HRS participants completed the Health Care and Nutrition Study, a sub-study which contains a dietary assessment[27], followed by two cognitive assessments in 2016 and 2018. The Framingham Heart Study is a community-based cohort study commenced in 1948 [28]. In 1971, children of the original cohort and their spouses formed the FOS cohort, who completed dietary assessments in examination cycles 5 (1991–1995), 6 (1995-1998) and 7 (1998–2001) and have undergone continuous surveillance of dementia through 2018.

We included participants with a baseline age >=45 years who completed >=1 valid Food Frequency Questionnaire (FFQ, being valid if total energy intake in 500-4500 kcal). Participants with dementia prior or at baseline were excluded from analyses (**Figure 1**). Analyses included 18163 participants, including 8360 from WII, 6758 from HRS and 3045 from FOS.

**Figure 1.**
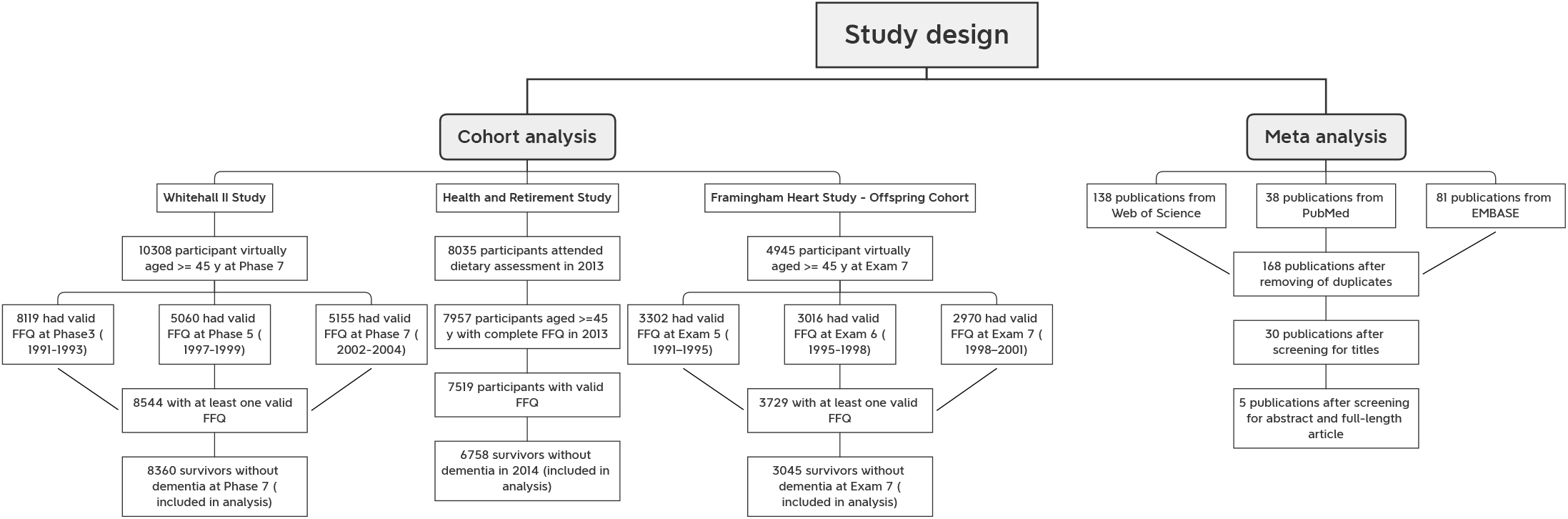
Study design

#### Mediterranean-Dietary Approaches to Stop Hypertension (MIND) diet score

In the three cohorts, participants were asked how often on average they consumed a specified amount of food in the past year using FFQs[23,25,28]. Despite of some cohort-specific modifications, the FFQs in all three cohort studies were based on the FFQ devised by Willett et al[29,30].

The primary exposure was the MIND diet score, which consisted of 10 recommended food groups and 5 restricted food groups [17] (**Supplemental Table 1**). Olive oil was scored 1 if it is used as the primary oil at home and 0 otherwise. For other recommended food groups, high consumption was assigned a concordance score of 1, moderate frequency was assigned 0.5, and low consumption was assigned 0. For restricted food groups, the scores were reversed. The total score was computed by summing over the 15 component scores (ranged: 0-15), with a higher score indicating higher adherence to the MIND diet. In the WII where information on olive oil was not available, the original score ranged 0-14 and was rescaled to 0-15. We took average MIND diet scores for participants with multiple dietary assessments before baseline to reduce within-person measurement error.

#### Ascertainment of incident all-cause dementia

The outcome of interest was incident all-cause dementia. The three cohort studies had different strategies in identifying incident dementia cases. In the WII study, dementia cases including subtypes were ascertained using linkage to 3 the HES database, the Mental Health Services Data Set, and the Office for National Statistics Mortality Register based on ICD-10 codes (F00-F03, F05.1, G30, and G31) though 2015[16]. Date of dementia was defined at the first record of first occurrence in all 3 databases. The HRS did not directly assess the status of dementia, and we adopted the Langa-Weir Classification strategy with both high specificity and sensitivity. Briefly, a participant was defined as dementia case if she or he had a poor objective performance in self interview or had a high score in cognitive disability assessed by a proxy respondent[31,32]. In the FOS, a dementia review panel consisting of neurologists and neuropsychologists reviewed every possible case of dementia according to criteria from the Diagnostic and Statistical Manual of Mental Disorders, 4th Edition (DSM-IV)[33]. After the decease of a participant, the panel reviews medical and nursing records to decide with the participant have had dementia since last examination[34].

#### Assessment of other covariates

We used baseline and historical questionnaires before baseline to collect the information of age (5-year groups), sex (male or female), education level (high school diploma or not), household income (only available in the HRS), frequency of vigorous physical activity (<1 times/month, 1-4 times/month, >1 times/week), current smoking status (yes or no), total energy intake, BMI categories (underweight or normal weight, overweight, or obesity), depressive symptom (yes or no), hypertension (yes or no), hypercholesterolemia (yes or no, not available in the HRS), diabetes (yes or no), stroke (yes or no), and cardiovascular diseases (yes or no). Total energy was calculated from the FFQ. Alcohol intake is not considered a confounding variable because wine is a component of the MIND diet. Detailed descriptions of design and validation of the questionnaires were shown elsewhere.

The assessment of some health conditions varied by cohorts. Depressive symptom was assessed using the Center for Epidemiologic Studies Depression Scale-8 (CES-D 8) for HRS participants, and participants scoring 4/8 or above were defined as having depressive symptom[35]. In the FOS and WII, a 20-item CES-D was used, and a score of 16/60 or above indicated depressive symptom[36,37]. In the FOS and WII where physical and blood examination was regularly administered, we additionally identified participants with hypertension [38](with anti-hypertensive medication or systolic blood pressure >=130 mmHg or diastolic blood pressure>=80 mmHg), diabetes [39](with oral hypoglycemic medication or insulin use, fasting blood glucose concentrations ≥126 mg/dL, or non-fasting blood glucose concentrations ≥200 mg/dL), and hypercholesterolemia [40](with cholesterol-lowering medication or total cholesterol concentrations ≥200 mg/dL, i.e., 5.20 mmol/L).

#### Statistical analyses

We calculated person-time from the study baseline (date of the latest wave of dietary assessment) to date of incident dementia, death of other causes, loss to follow-up, or the end of follow-up (2015 for WII, 2018 for FOS and HRS), whichever occurred first.

In the primary analyses, we assessed the relations of MIND diet score and its components (average levels in repeated assessments) to incident dementia with Cox proportional hazard models, with adjustments for baseline sociodemographic factors, health-related behaviors, and health conditions, including age (5-year groups), sex (male or female), total energy intake, education level (high school diploma or not), household income (only in HRS), frequency of vigorous physical activity (<1 times/month, 1-4 times/month, >1 times/week), current smoking status (yes, no), BMI categories (underweight or normal weight, overweight, and obesity), hypertension (yes or no), diabetes (yes or no), stroke (yes or no), hypercholesterolemia (yes or no, not available in HRS), and cardiovascular diseases (yes or no). MIND diet was used as tertile-defined categorical and continuous (per 3-unit increment) variables. The linearity assumption of the model for continuous score was evaluated by comparing the Akaike information criterion of a linear model with a quadratic model. Analyses were conducted within each cohort separately, and the results were pooled from the three cohorts using random-effect models. We performed prespecified stratified analyses by age (<70 y or >=70 y), sex (female or male), current smoking status (yes or no), and BMI categories (Underweight or normal weight, or overweight or obesity). We tested for potential interactions for these variables using likelihood ratio tests.

In the exploratory analysis, we further assessed the temporal relations of the MIND diet score to all-cause dementia, comparing the recent MIND score and the remote MIND in relations to dementia in studies with repeated dietary assessments (WII and FOS). In the WII where participants received dietary assessments in 1991-1993, 1997-1999, and 2002-2004, remote MIND score was calculated from FFQ in 1991-1993, and recent MIND score calculated from that in 2002-2004. Similarly, in the FOS, we calculated the remote and recent MIND score in 1991–1995 and 1998–2001, respectively. The study baselines were defined as the date of the latest wave of dietary assessment (e.g., 2002-2004 in WII) in the temporal analysis. In addition, we assessed the combined associations of the remote and recent MIND score with incident all-cause dementia in later life.

We conducted several sensitivity analyses to test the robustness of our primary findings. First, we additionally adjusted the models for depression status, because although depression itself could be a confounding factor related to both dietary pattern and dementia, it might be an important mediator[41]. Given the long preclinical period of dementia, dietary changes might be a preclinical sign of dementia, so we first excluded participants who developed dementia with the first two years of follow-up to further reduce reverse causation. We alternatively used pre-specified cutoffs (7 and 9 points) of MIND diet score rather than the distribution-driven cut-offs to test the robustness of the association. We restricted our analyses in participants without baseline stroke to reduce the potential confounding effect of stroke. Stroke itself is related to higher risk of later life dementia and might introduce dietary changes, but could also be a potential mediator in the pathway from diet to dementia[42], so we only excluded stroke survivors as a sensitivity analysis. Furthermore, we adjusted the models for continuous age rather than age groups to account for potential residual confounding where applicable.

We reported two-sided P-values throughout and a P-values less than 0.05 was considered an indicator for statistical significance. Statistical analyses were performed using SAS version 9.4 (SAS Institute, Cary, NC).

### 2. Meta-analysis

We conducted meta-analysis using data from this study and previous full-length publications reporting cohort studies that assessed the relations of MIND diet to incident dementia or AD in the general middle-aged and older adults. Following the preferred reporting items for systematic reviews and meta-analyses (PRISMA) guidelines, we registered the protocol on INPLASY (INPLASY202270127). We systematically searched PubMed, Embase, and Web of Science on July 26, 2022 using strategy presented in **Supplemental table 2**, and we additionally screened the references of the literature.

We included cohort studies that assessed the associations of MIND diet and incident dementia or AD and extracted the name of the first author, publication year, cohort name, country, number of participants, age range at baseline, dietary assessment methods, and dementia ascertainment methods, risk estimates and 95% CI from the multivariable-adjusted models and included covariates. We used the Newcastle-Ottawa scale[43] to assess the risk of bias in included studies. Two authors (HC and LH) independently screened the literature, extracted the data, and assessed the risk of bias. Disagreement and discordance were resolved by consensus between the two authors (**Figure 1**).

We used random-effect models to pool the risk estimates comparing extreme MIND diet categories and reported the I^2^ statistic. An I^2^ statistic <=40% indicated unimportant heterogeneity; I^2^ statistic in 30-60% represents moderate heterogeneity; and I^2^ statistic over 50% represents substantial heterogeneity[44]. Publication bias was tested by using Egger’s test. The analysis were performed using R package ‘meta’[45].

### 3. Patient and public involvement

No patient or participant was involved in the conceptualization, design, and implementation of the current study.

## Results

### Cohort analyses

In the cohort analyses, we included a total of 18,163 women and men who were free of dementia at baseline, including 8,360 participants (30.9% female) from WII, 6,758 participants (58.7% female) from HRS, and 3,045 participants (54.6% female) from FOS (**Table 1**). The means (SDs) of baseline MIND diet scores were 8.3 (1.4) in WII, 7.1 (1.9) in HRS, and 8.1 (1.6) in FOS (**Supplemental Table 3**).

**Table 1.**
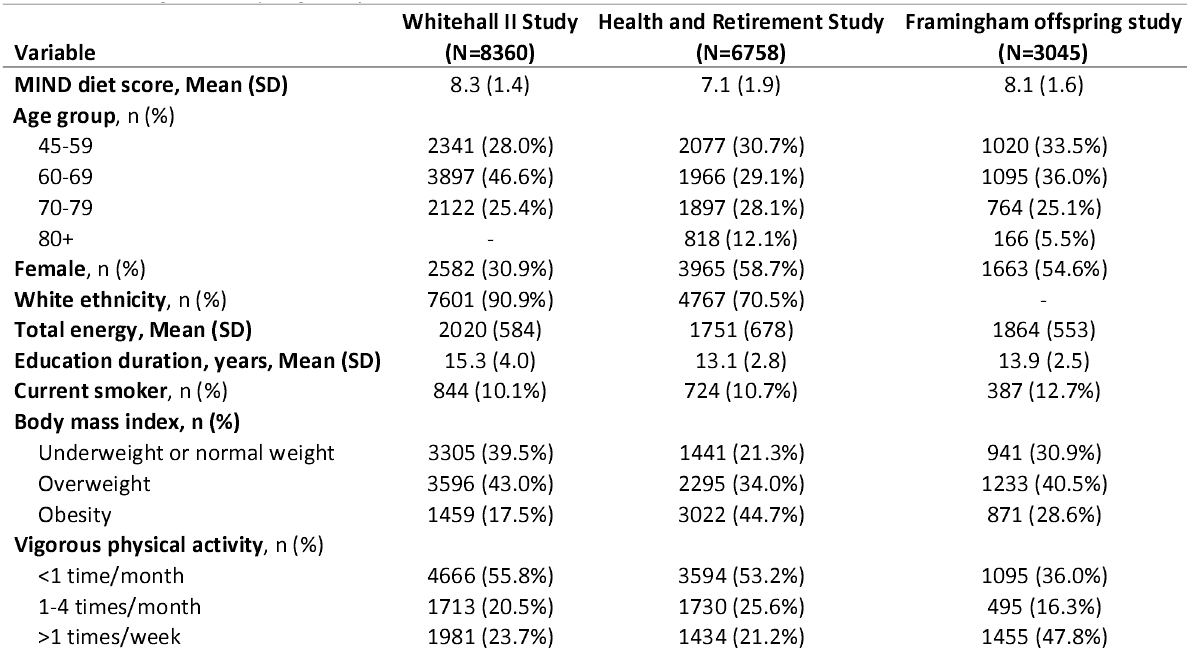
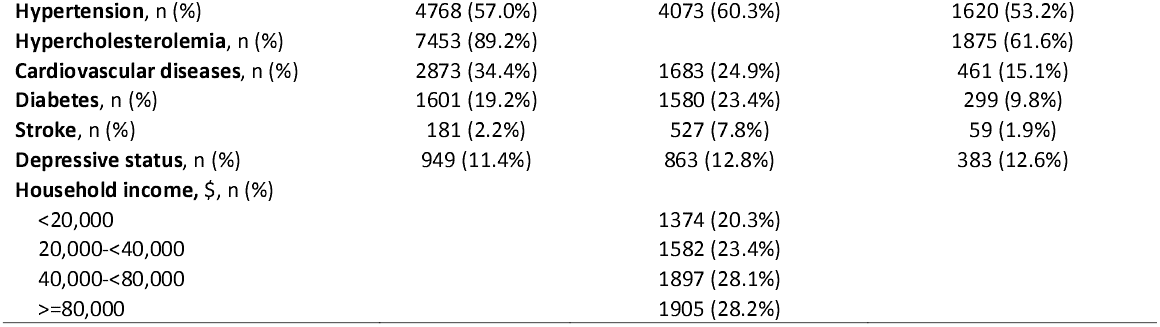
Baseline characteristics of 8360 participants in the Whitehall II study, 6758 participants in the Health and Retirement Study, and 3045 participants in the Framingham Offspring Study

| **Variable** | **Whitehall II Study**<br>**(N=8360)** | **Health and Retirement Study**<br>**(N=6758)** | **Framingham offspring study**<br>**(N=3045)** |
| --- | --- | --- | --- |
| **MIND diet score, Mean (SD)** | 8.3 (1.4) | 7.1 (1.9) | 8.1 (1.6) |
| **Age group, n (%)** |  |  |  |
| 45-59 | 2341 (28.0%) | 2077 (30.7%) | 1020 (33.5%) |
| 60-69 | 3897 (46.6%) | 1966 (29.1%) | 1095 (36.0%) |
| 70-79 | 2122 (25.4%) | 1897 (28.1%) | 764 (25.1%) |
| 80+ | - | 818 (12.1%) | 166 (5.5%) |
| **Female, n (%)** | 2582 (30.9%) | 3965 (58.7%) | 1663 (54.6%) |
| **White ethnicity, n (%)** | 7601 (90.9%) | 4767 (70.5%) | - |
| **Total energy, Mean (SD)** | 2020 (584) | 1751 (678) | 1864 (553) |
| **Education duration, years, Mean (SD)** | 15.3 (4.0) | 13.1 (2.8) | 13.9 (2.5) |
| **Current smoker, n (%)** | 844 (10.1%) | 724 (10.7%) | 387 (12.7%) |
| **Body mass index, n (%)** |  |  |  |
| Underweight or normal weight | 3305 (39.5%) | 1441 (21.3%) | 941 (30.9%) |
| Overweight | 3596 (43.0%) | 2295 (34.0%) | 1233 (40.5%) |
| Obesity | 1459 (17.5%) | 3022 (44.7%) | 871 (28.6%) |
| **Vigorous physical activity, n (%)** |  |  |  |
| <1 time/month | 4666 (55.8%) | 3594 (53.2%) | 1095 (36.0%) |
| 1-4 times/month | 1713 (20.5%) | 1730 (25.6%) | 495 (16.3%) |
| >1 times/week | 1981 (23.7%) | 1434 (21.2%) | 1455 (47.8%) |
| **Hypertension, n (%)** | 4768 (57.0%) | 4073 (60.3%) | 1620 (53.2%) |
| **Hypercholesterolemia, n (%)** | 7453 (89.2%) |  | 1875 (61.6%) |
| **Cardiovascular diseases, n (%)** | 2873 (34.4%) | 1683 (24.9%) | 461 (15.1%) |
| **Diabetes, n (%)** | 1601 (19.2%) | 1580 (23.4%) | 299 (9.8%) |
| **Stroke, n (%)** | 181 (2.2%) | 527 (7.8%) | 59 (1.9%) |
| **Depressive status, n (%)** | 949 (11.4%) | 863 (12.8%) | 383 (12.6%) |
| **Household income, \$, n (%)** | | | |
| <20,000 |  | 1374 (20.3%) |  |
| 20,000-<40,000 |  | 1582 (23.4%) |  |
| 40,000-<80,000 |  | 1897 (28.1%) |  |
| >=80,000 |  | 1905 (28.2%) |  |

Over 171,079 person-years, 802 participants (222/110,352 person-years in WII, 338/29,182 person-years in HRS, and 242/31,658 person-years in FOS) developed incident all-cause dementia. In the multivariable-adjusted model (**Table 2**), every 3-point increment was related to a 20% lower risk of dementia (pooled hazard ratio (HR): 0.80, 95% confidence interval (CI): 0.70-0.91, P-trend=0.0014, P-heterogeneity=0.3039). The HRs (95% CIs) were 0.92 (0.69-1.24) in WII, 0.82 (0.68-0.99) in HRS, and 0.68 (0.52-0.88) in FOS. Potential contributors included green-leafy vegetables, other vegetables, berries, nuts, olive oil, and beans (**Supplemental Table 4**). For example, the HRs were for green-leafy vegetables (>=6 v. <=2 servings/wk) 0.53 (0.36-0.80) in FOS, and other vegetables (>=7 v. <5 servings/wk) showed significant protective association in HRS (0.66; 0.51-0.85).

**Table 2.**
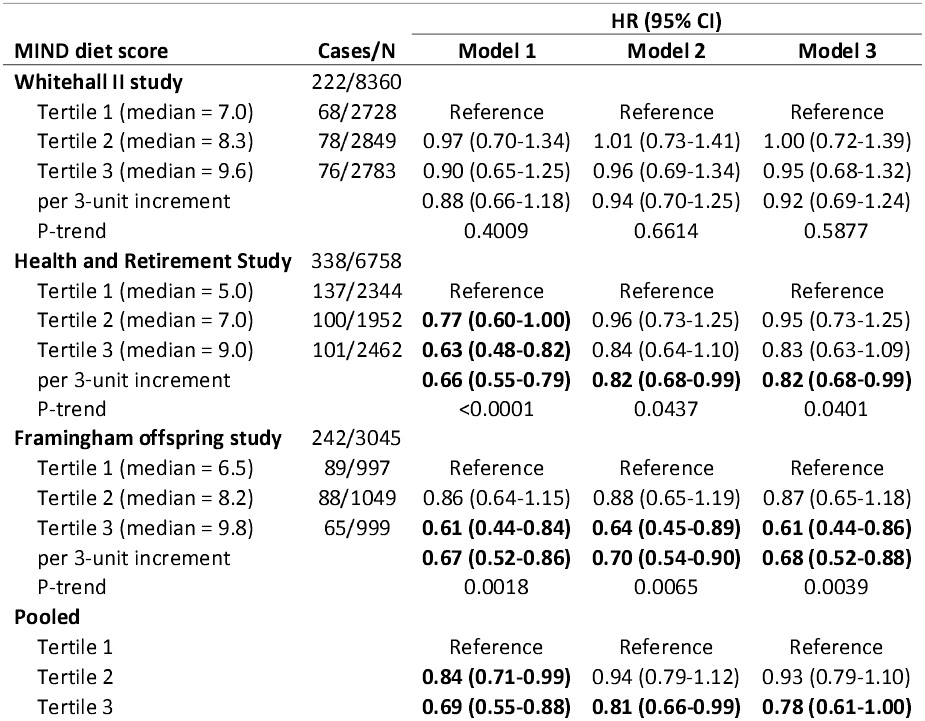
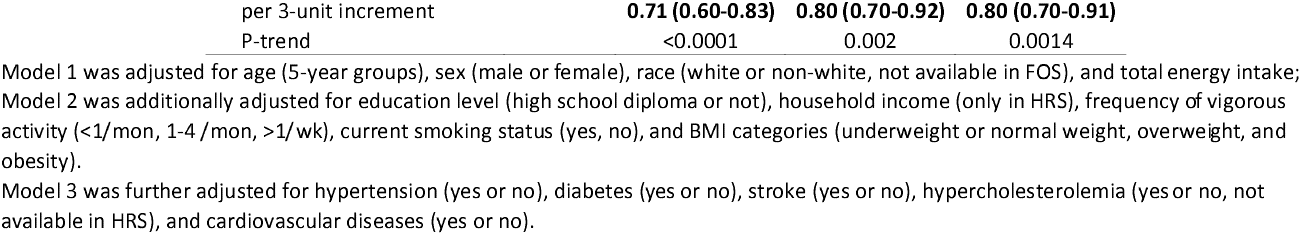
Multivariable adjusted hazard ratios (HRs) and 95% confidence intervals (CIs) for incident dementia according to tertiles of Mediterranean-DASH Intervention for Neurodegenerative Delay (MIND) diet score

In the temporal analysis (**Figure 2 and Supplemental Table 5**) among 4818 WII participants, the HRs (95% CIs) was 1.04 (0.71-1.52) for every 3-point increment in the remote MIND score, and 0.76 (0.53-1.09) for recent MIND score. Change from high to low MIND diet score was associated with 168% (23%-484%) increased risk of dementia, compared with those who had low score in both assessments. In 2347 FOS participants, the HRs were 0.66 (0.50-0.86) and 0.67 (0.51-0.88) for remote and recent MIND score, respectively. High MIND diet score in both assessments was related to 56% (23%-74%) lower hazard of dementia, compared with those who had low score in both assessments.

**Figure 2.**
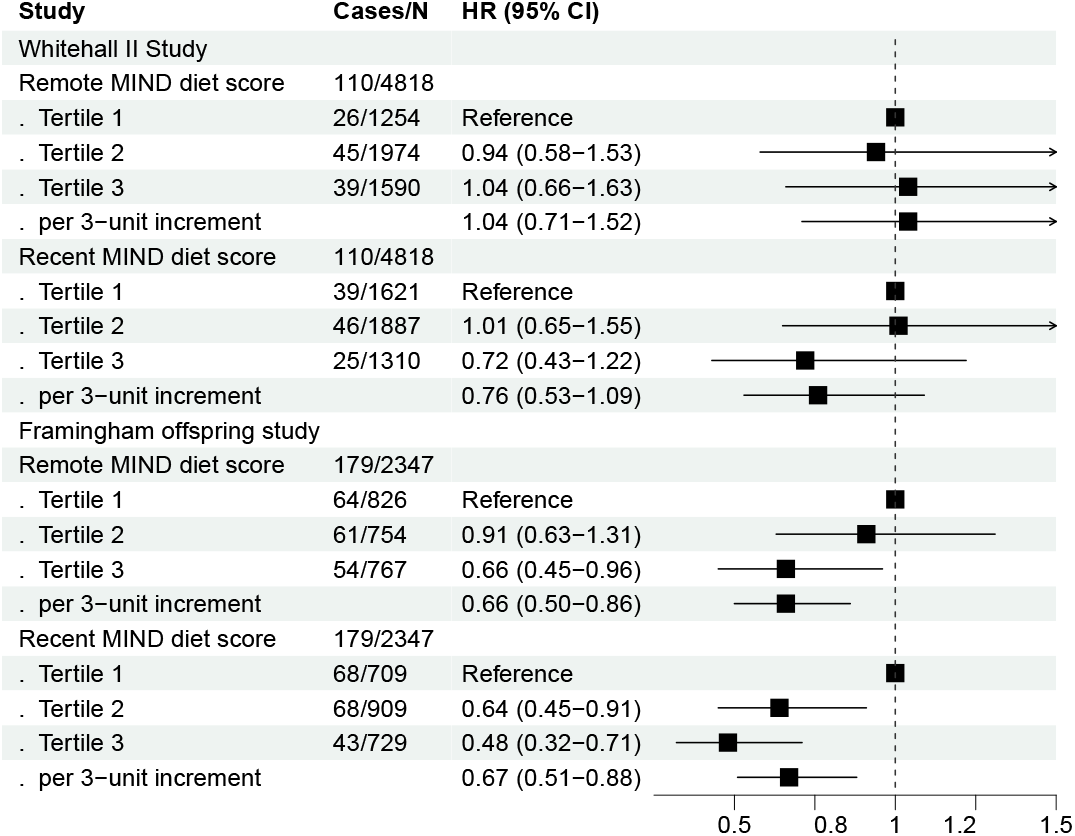
Multivariable adjusted hazard ratios (95% confidence intervals) for incident dementia according to recent and remote Mediterranean-DASH Intervention for Neurodegenerative Delay (MIND) diet scores Cox proportional hazard models were adjusted for age (5-year groups), sex (male or female), total energy intake, education level (high school diploma or not), household income (only in HRS), physical activity (cohort-specific categories), current smoking status (yes, no), BMI categories (underweight or normal weight, overweight, and obesity), hypertension (yes or no), diabetes (yes or no), stroke (yes or no), hypercholesterolemia (yes or no, not available in HRS), and cardiovascular diseases (yes or no).

In the subgroup analyses (**Supplemental Table 6**), although no significant effect modification was observed (P-interactions>0.10 for all tests), we found null association in current smokers (1.03, 0.65-1.65), but in non-smokers the relation was strong and significant (0.76, 0.65-0.87). The relations did not vary significantly by sex, age, and BMI. For example, the HR (95% CI) was 0.75 (0.62-0.91) for female, no significantly lower that for male (0.88; 0.68-1.13).

The primary findings persisted in the sensitivity analyses (**Supplemental Table 7**). When using alternative cutoffs, the HR (95% CI) for a score of 7.0-8.9 was 0.91 (0.75-1.10), and 0.70 (0.55-0.89) for 9.0-15.0, compared with MIND score less than 7.0. When we further adjusted the models for depressive symptoms, every 3-point increment in MIND score was associated with 20% (95% CI: 8%-31%) decreased risk of dementia. When we excluded incident dementia cases within the first two years of follow-up, the pooled HR was 0.82 (0.72-0.95). When we excluded participants with a stroke history, the association remained strong (pooled HR: 0.82; 95% CI: 0.71-0.95). Adjusting for continuous age rather than categorical age did not substantially change the findings.

### Meta-analysis

We screened 258 studies and included eleven cohort findings from six eligible studies (including this study)[18,21,46–48]. Specifically, we included the two largely non-overlap sub-cohorts in the Rotterdam Study, and excluded the analyses on Memory and Aging Project in the study of Vu et al. because it included same sample as in the study of Morris et al. The characteristics of the included studies were shown in **Supplemental table 8. Supplemental table 9** presents the assessments of risk of bias using the Newcastle-Ottawa scale. All studies obtained a score of at least seven (low risk of bias).

Pooling 11 risk estimates of 224,140 participants, the highest category of MIND diet score was related to 18% lower risk of dementia (pooled relative risk 0.82, 95% CI: 0.74-0.92, Figure 3), with substantial heterogeneity (I^2^=74%, P-heterogeneity=<0.01). We did not observe significant publication bias (Egger’s P= 0.096). The funnel plot was shown in **Supplemental figure 1**. No single study substantially affected the pooled results (data not shown).

**Figure 3.**
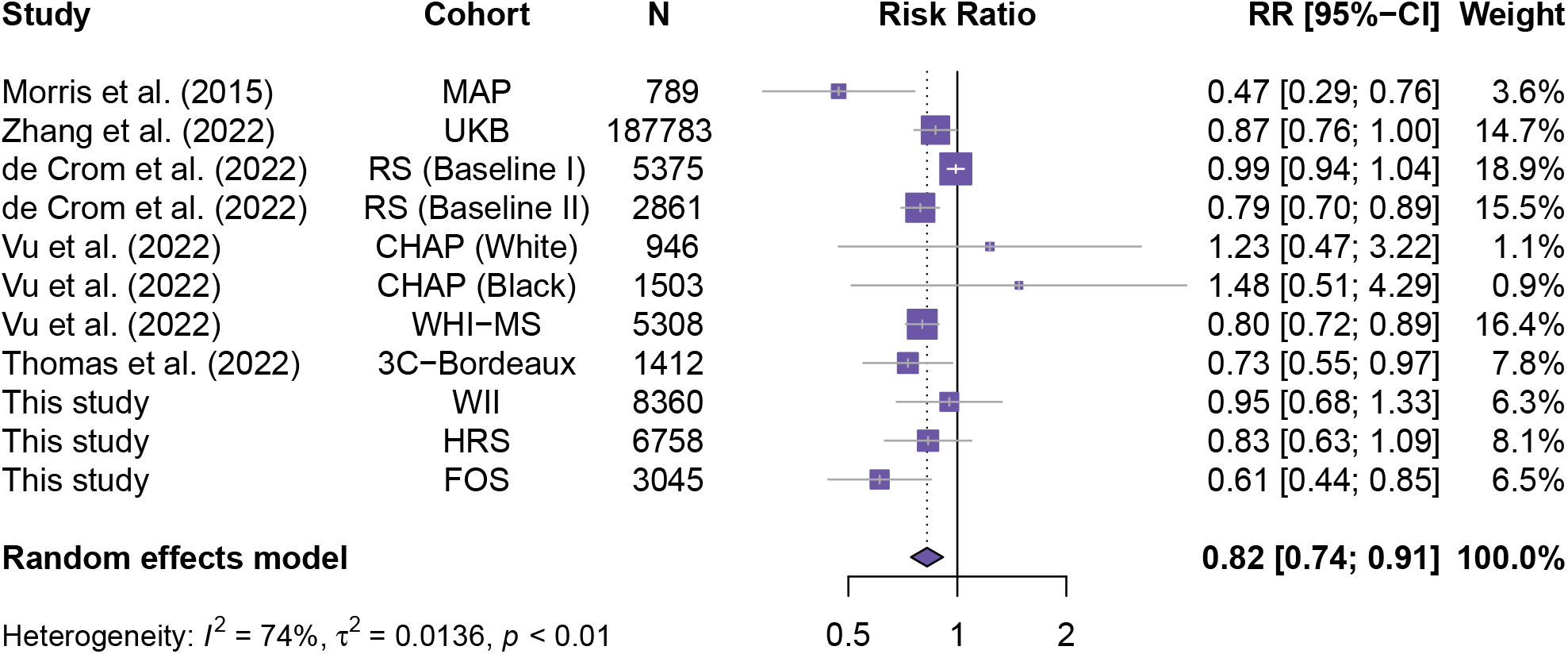
Pooled risk ratio for incident dementia comparing the highest and lowest groups of MIND diet score MIND: Mediterranean-DASH Intervention for Neurodegenerative Delay, MAP: Memory and Aging Project, UKB: UK Biobank, RS: Rotterdam Study, CHAP: Chicago Health and Aging Project, WHI-MS: Women’s Health Initiative Memory Study, 3C: Three-City Study, WII: Whitehall II Study, HRS: Health and Retirement Study, FOS: Framingham Offspring Study

## Discussion

### Principal findings

In the current study, higher adherence to the MIND diet was related to lower risk of incident all-cause dementia. In an exploratory temporal analysis, we observed that in the WII study only recent MIND diet showed an inverse association, while both remote and recent MIND diet demonstrated strong associations in the FOS. In the meta-analysis, we observed that MIND diet was associated with lower risk of dementia, although previous evidence was substantially heterogeneous. Future observational and interventional studies are needed to assess the associations across different cultural backgrounds.

### Comparison with other studies

Until now, only several cohort studies have associated MIND diet with incident all-cause dementia, although it has been linked to cognitive function and its decline[17,19]. In a 4.5-years study among 923 US adults, higher MIND diet score was related to substantially lower risk of incident all-cause dementia [18](HR: 0.47, 95% CI: 0.29-0.76 comparing extreme tertiles). A recent study in the Netherlands reported a significant protective association of the MIND diet over the first 7 years of follow-up (0.85, 0.74-0.98 per SD increment), but such association were no longer significant in longer-term follow-ups[21]. Based on these findings, we utilized three well-characterized cohort studies and confirmed the potentially protective associations of MIND diet.

In the WII where the overall association was non-significant, recent MIND diet showed a direction of protective association, while the remote diet score showed null association. In particular, a subgroup of participants who changed from high (remote years) to low (recent years) MIND diet score had a significant higher risk of later-life dementia. In the FOS, the relations were significantly inverse for both remote and recent MIND diet. To date, most studies evaluating the link between MIND diet and other cognitive outcomes (including cognitive function, its decline and incident cognitive impairment) are limited by small sample sizes or short-term follow-ups, which might suffer from reverse causation induced by early behavioral changes resulting from preclinical dementia. Our study provided valuable evidence with relatively long-term follow-up (5 to 17 years), but still cannot fully eliminate reverse causation and limited power in the analysis of changes in MIND score.

MIND diet emphasizes several brain health food groups, such as green leafy vegetables, olive oil, and berries[49], which are confirmed in this study. These food groups are believed to confer protection against brain aging through their antioxidant and anti-inflammatory properties and inhibition of b-amyloid deposition of specific nutrients, such as vitamin E[50], folate[12], flavonoids[11], and carotenoids[9]. Currently, a 3-year, multicenter, randomized controlled trial of the MIND diet on cognitive decline is being conducted in US adults, which is expected to provide valuable evidence for dietary strategies in dementia prevention[49].

Interestingly, we observed substantial heterogeneity in current literature. The differences in education levels, age ranges, and duration of follow-ups among cohorts might be the underlying reasons. For example, in the WII cohort where participants were relatively better-educated civil servants, a previous study have pointed out that midlife alternate healthy eating index was not related to later-life dementia[16]. In the Rotterdam Study (Baseline I) cohort, the association was significant in the first 7-year follow-up, but was also attenuated to null in the 15.6-year follow-up [21], implying that the long-term association might still be unclear. Also, the cultural backgrounds in different populations might be an important underlying source of heterogeneity, and current literature on the MIND diet is largely based on western population. Therefore, whether this pattern is beneficial for primary prevention of dementia in other races warrants exploration, and evidence for specific components is even more limited.

### Strengths and limitations of study

The strengths of the current study included long-term follow-up in population-based cohort studies with validated approaches to assess dietary intake. Nevertheless, our findings should be interpreted with caution due to some limitations. First, we were unable to specifically assess the relations of MIND diet to subtypes of dementia because of the inconsistent definitions across cohorts. Second, measurement errors are inevitable, and confounding owing to unmeasured food items could still lead to biased estimation of the relation of MIND diet to dementia. Therefore, our observational findings of this study do not necessarily reflect causal relations. Third, the cohort studies and meta-analysis mainly consisted of western population, so our findings might not be generalizable to populations with different cultural and ethnical backgrounds. Furthermore, misclassification of dementia cases might still be an issue in the current study, especially in the cohorts relying on linkage to data with health electronic systems (e.g., WII) and those based on simplified cognitive tests (e.g., HRS). Finally, consumption of some food groups, such as olive oils, might not be popular in specific countries at the study baseline, and further investigations of these food groups is warranted.

### Conclusion and public health implications

Adherence to the MIND diet is prospectively related to dementia, although heterogeneity was observed across studies. Future studies are needed to develop and refine the specific MIND diet for different populations and to identify the optimal intervention time window for dementia prevention.

## Supporting information

STROBE checklist

## Data Availability

Data are available on request for bone fide investigators from managing institutions of the three cohort studies: https://www.ucl.ac.uk/epidemiology-health-care/research/epidemiology-and-public-health/research/whitehall-ii for the WII, https://www.framinghamheartstudy.org/fhs-for-researchers/ for the FOS, and https://hrs.isr.umich.edu for the HRS. Data used in the meta-analysis are shown in the supplemental table.

## Statements

### Ethical approval

All participants provided informed consent to be eligible to participate in the cohort studies. The WII was approved by University College London Medical School committee, the HRS was approved by University of Michigan Institutional Review Board (IRB), and the FOS was approved by Boston University Medical Center IRB.

### Data availability statement

Data are available on request for bone fide investigators from managing institutions of the three cohort studies:

https://www.ucl.ac.uk/epidemiology-health-care/research/epidemiology-and-public-health/research/whitehall-ii for the WII, https://www.framinghamheartstudy.org/fhs-for-researchers/ for the FOS, and https://hrs.isr.umich.edu for the HRS. Data used in the meta-analysis are shown in the supplemental table.

## Acknowledgements

HC and CY designed and conceptualized the study, interpreted the findings, and drafted the manuscript. KD, YZ, XL, DML, AA, and WW revised the manuscript. HC performed cohort analyses. HC and LH independently conducted literature screening and data extraction for meta-analysis. YH, LH and YT provided assistance in data analysis and interpretation of the findings. DPUK provided data access for this project through MRC grant ref MR/L023784/2.

## Dissemination to participants and related patient and public communities

We plan to disseminate our findings to the general public in a press release.

