## Supplemental Information for "Mediterranean-DASH Intervention for Neurodegenerative Delay diet and risk of dementia: three prospective studies and meta-analysis of cohort studies"

### Supplementary materials

**Supplemental Table 1**. Scoring system of the Mediterranean-DASH Intervention for Neurodegenerative Delay (MIND) diet score

**Supplemental Table 2.** Searching strategy in the meta-analysis

**Supplemental Table 3.** Mean and standard deviation (SD) of the Mediterranean-DASH Intervention for Neurodegenerative Delay (MIND) diet

**Supplemental Table 4.** Multivariable adjusted hazard ratios (95% confidence intervals) for incident dementia according to remote, recent, and change from remote to recent Mediterranean-DASH Intervention for Neurodegenerative Delay (MIND) diet scores

**Supplemental Table 5.** Multivariable adjusted hazard ratios (95% confidence intervals) for incident dementia according to tertiles of Mediterranean-DASH Intervention for Neurodegenerative Delay (MIND) diet score by subgroups of participants

**Supplemental Table 6.** Multivariable adjusted hazard ratios (95% confidence intervals) for incident dementia according to components of Mediterranean-DASH Intervention for Neurodegenerative Delay (MIND) diet score

**Supplemental Table 7**. Multivariable adjusted hazard ratios (95% confidence intervals) for incident dementia according of Mediterranean-DASH Intervention for Neurodegenerative Delay (MIND) diet score in sensitivity analyses

**Supplemental Table 8**. Characteristics of included studies in the meta-analysis

**Supplemental Table 9**. Risk of bias of the included studies in the Newcastle-Ottawa scale

**Supplemental Figure 1**. Funnel plot in the meta-analysis

**Supplemental Table 1**. Scoring system of the Mediterranean-DASH Intervention for Neurodegenerative Delay (MIND) diet score

| **Food group** | **0 unit** | **0.5 unit** | **1 unit** |
| --- | --- | --- | --- |
| Green-leafy vegetables | <=2 servings/wk | >2-<6 servings/wk | >=6 servings/wk |
| Other vegetables | <5 servings/wk | 5-<7 servings/wk | >=7 servings/wk |
| Berries | <1 serving/wk | 1 serving/wk | >=6 servings/wk |
| Nuts | <1 serving/mo | 1 serving/mo-<5 servings/wk | >=6 servings/wk |
| Olive oil | Not primary oil | - | Primary oil |
| Butter and Margarine | >2 T/d | 1-2 T/d | <1 T/d |
| Cheese | >=7 servings/wk | 1-6 servings/wk | <1 servings/wk |
| Whole grains | <1 serving/d | 1-2 servings/d | <1 serving/wk |
| Fish (not fried) | <1 meal/mon | 1-3 meals/mon | >=1 meals/wk |
| Beans | <1 meal/wk | 1-3 meals/wk | >3 meals/wk |
| Poultry (not fried) | <1 meal/wk | 1 meal/wk | >=2 meals/wk |
| Red meat and products | >=7 servings/wk | 4-6 meals/wk | <4 meals/wk |
| Fast fried foods | >=4 times/wk | 1-3 times/wk | <1 time/wk |
| Pastries and sweets | >=7 servings/wk | 5-6 servings/wk | <5 servings/wk |
| Wine | >1 glass/d or never | 1/mo-6/wk | 1 glass/d |

**Supplemental Table 2.** Searching strategy in the meta-analysis

| **Database** | **Searching strategy** | **N** |
| --- | --- | --- |
| PubMed | (((("MIND diet"[Title/Abstract]) OR ("Mediterranean-DASH Diet Intervention for Neurodegenerative Delay"[Title/Abstract]) OR ("Mediterranean-Dietary Approach to Systolic Hypertension (DASH) diet intervention for neurodegenerative delay"[Title/Abstract]))) AND ((dementia [Title/Abstract]) OR (“alzheimer disease” [MeSH Terms])) | 38 |
| WebOfScience | (TS=(MIND diet) OR TS=(Mediterranean-DASH Diet Intervention for Neurodegenerative Delay)) AND (TS=(dementia) OR TS=(alzheimer’s disease)) | 138 |
| EMBASE | ('mind diet'/exp OR 'Mediterranean-dash diet intervention for neurodegenerative delay' OR 'Mediterranean-dietary approach to systolic hypertension (dash) diet intervention for neurodegenerative delay') AND (‘alzheimer disease’ OR dementia) | 81 |

**Supplemental Table 3.** Mean and standard deviation (SD) of the Mediterranean-DASH Intervention for Neurodegenerative Delay (MIND) diet

|  | **Whitehall II Study (N=8360)** | | | **Health and Retirement Study (N=6758)** | **Framingham offspring study (N=3045)** | | |
| --- | --- | --- | --- | --- | --- | --- | --- |
| **Component** | Average | Recent | Remote |  | Average | Recent | Remote |
| **Total** | 8.3 (1.40) | 8.5 (1.60) | 8.2 (1.56) | 7.1 (1.90) | 8.1 (1.61) | 8.3 (1.83) | 7.9 (1.77) |
| Green-leafy vegetables | 0.5 (0.33) | 0.5 (0.38) | 0.5 (0.38) | 0.4 (0.39) | 0.5 (0.33) | 0.5 (0.40) | 0.5 (0.39) |
| Other vegetables | 0.9 (0.24) | 0.9 (0.22) | 0.9 (0.30) | 0.6 (0.44) | 0.7 (0.33) | 0.7 (0.39) | 0.7 (0.39) |
| Berries | 0.4 (0.37) | 0.4 (0.43) | 0.4 (0.42) | 0.3 (0.42) | 0.3 (0.29) | 0.3 (0.37) | 0.3 (0.35) |
| Nuts | 0.3 (0.25) | 0.3 (0.30) | 0.3 (0.28) | 0.4 (0.34) | 0.3 (0.23) | 0.3 (0.29) | 0.3 (0.27) |
| Olive oil |  |  |  | 0.3 (0.45) | 0.3 (0.38) | 0.4 (0.49) | 0.2 (0.42) |
| Butter and Margarine | 0.6 (0.38) | 0.7 (0.44) | 0.6 (0.46) | 0.9 (0.26) | 0.8 (0.33) | 0.8 (0.38) | 0.7 (0.42) |
| Cheese | 0.5 (0.23) | 0.5 (0.28) | 0.5 (0.26) | 0.5 (0.29) | 0.5 (0.23) | 0.5 (0.28) | 0.5 (0.28) |
| Whole grains | 0.3 (0.26) | 0.3 (0.30) | 0.3 (0.32) | 0.3 (0.31) | 0.3 (0.27) | 0.3 (0.32) | 0.3 (0.33) |
| Fish (not fried) | 0.9 (0.23) | 0.9 (0.28) | 0.9 (0.27) | 0.6 (0.41) | 0.9 (0.21) | 0.9 (0.25) | 0.9 (0.24) |
| Beans | 0.8 (0.25) | 0.8 (0.29) | 0.8 (0.29) | 0.4 (0.35) | 0.6 (0.25) | 0.6 (0.30) | 0.6 (0.30) |
| Poultry (not fried) | 0.6 (0.34) | 0.6 (0.39) | 0.5 (0.39) | 0.2 (0.37) | 0.8 (0.26) | 0.8 (0.32) | 0.8 (0.31) |
| Red meat and products | 0.5 (0.37) | 0.5 (0.42) | 0.5 (0.42) | 0.6 (0.44) | 0.6 (0.36) | 0.6 (0.43) | 0.6 (0.43) |
| Fast fried foods | 0.8 (0.21) | 0.8 (0.28) | 0.9 (0.21) | 0.9 (0.22) | 0.9 (0.17) | 0.9 (0.21) | 0.9 (0.22) |
| Pastries and sweets | 0.2 (0.34) | 0.3 (0.41) | 0.2 (0.37) | 0.5 (0.48) | 0.4 (0.38) | 0.4 (0.46) | 0.4 (0.45) |
| Wine | 0.4 (0.25) | 0.4 (0.32) | 0.4 (0.28) | 0.1 (0.23) | 0.3 (0.23) | 0.3 (0.28) | 0.3 (0.27) |

Olive oil was not measured in the Whitehall II FFQ.

**Supplemental Table 4.** Multivariable adjusted hazard ratios (95% confidence intervals) for incident dementia according to components of Mediterranean-DASH Intervention for Neurodegenerative Delay (MIND) diet score

| **Component (High vs. low)** | **Whitehall II Study (N=8360)** | **Health and Retirement Study (N=6758)** | **Framingham offspring study (N=3045)** |
| --- | --- | --- | --- |
| Green-leafy vegetables | 0.80 (0.53-1.20) | 0.92 (0.68-1.25) | 0.53 (0.36-0.80) |
| Other vegetables | 0.93 (0.52-1.66) | 0.66 (0.51-0.85) | 0.68 (0.45-1.02) |
| Berries | 0.76 (0.53-1.11) | 0.86 (0.65-1.13) | 0.84 (0.52-1.37) |
| Nuts | 0.74 (0.43-1.28) | 0.62 (0.43-0.88) | 0.81 (0.45-1.46) |
| Olive oil |  | 0.93 (0.71-1.22) | 0.51 (0.35-0.74) |
| Butter and Margarine | 1.11 (0.76-1.63) | 0.70 (0.45-1.11) | 1.28 (0.84-1.92) |
| Cheese | 1.13 (0.62-2.05) | 1.60 (1.01-2.54) | 0.81 (0.45-1.46) |
| Whole grains | 0.91 (0.54-1.54) | 0.69 (0.48-1.00) | 1.15 (0.69-1.91) |
| Fish (not fried) | 0.88 (0.48-1.62) | 0.99 (0.75-1.31) | 0.75 (0.40-1.39) |
| Beans | 0.82 (0.47-1.44) | 0.90 (0.66-1.24) | 0.55 (0.31-0.97) |
| Poultry (not fried) | 0.87 (0.59-1.31) | 1.01 (0.75-1.35) | 1.00 (0.62-1.58) |
| Red meat and products | 0.91 (0.62-1.35) | 1.36 (1.04-1.81) | 1.44 (0.95-2.22) |
| Fast fried foods | 0.89 (0.44-1.78) | 1.13 (0.69-1.85) | 0.60 (0.21-1.69) |
| Pastries and sweets | 0.63 (0.42-0.94) | 0.90 (0.70-1.14) | 0.84 (0.58-1.21) |
| Wine (moderate vs. non-moderate) | 0.80 (0.48-1.35) | 1.11 (0.66-1.87) | 1.11 (0.64-1.94) |

Cox proportional hazard models were adjusted for age (5-year groups), sex (male or female), total energy intake, education level (high school diploma or not), household income (only in HRS), physical activity (cohort-specific categories), current smoking status (yes, no), BMI categories (underweight or normal weight, overweight, and obesity), hypertension (yes or no), diabetes (yes or no), stroke (yes or no), hypercholesterolemia (yes or no, not available in HRS), and cardiovascular diseases (yes or no).

**Supplemental Table 5.** Multivariable adjusted hazard ratios (95% confidence intervals) for incident dementia according to remote, recent, and change from remote to recent Mediterranean-DASH Intervention for Neurodegenerative Delay (MIND) diet scores

|  | **Whitehall II Study** | | | **Framingham offspring study** | | |
| --- | --- | --- | --- | --- | --- | --- |
|  | **Cases/N** | **HR (95% CI)** | **P-value** | **Cases/N** | **HR (95% CI)** | **P-value** |
| **Remote MIND diet score** | 110/4818 |  |  | 179/2347 |  |  |
| Tertile 1 | 38/1811 | Reference |  | 61/790 | Reference |  |
| Tertile 2 | 30/1364 | 0.94 (0.58-1.53) | 0.8149 | 60/760 | 0.91 (0.63-1.31) | 0.6082 |
| Tertile 3 | 42/1643 | 1.04 (0.66-1.63) | 0.8669 | 58/797 | **0.66 (0.45-0.96)** | 0.0308 |
| per 3-unit increment |  | 1.04 (0.71-1.52) | 0.8293 | 179/2347 | **0.66 (0.50-0.86)** | 0.0024 |
| **Recent MIND diet score** | 110/4818 |  |  | 179/2347 |  |  |
| Tertile 1 | 38/1811 | Reference |  | 65/679 | Reference |  |
| Tertile 2 | 30/1364 | 1.01 (0.65-1.55) | 0.9765 | 70/920 | **0.64 (0.45-0.91)** | 0.0124 |
| Tertile 3 | 42/1643 | 0.72 (0.43-1.22) | 0.2225 | 44/748 | **0.48 (0.32-0.71)** | 0.0003 |
| per 3-unit increment |  | 0.76 (0.53-1.09) | 0.1341 | 179/2347 | **0.67 (0.51-0.88)** | 0.0036 |
| **Change in MIND diet score** |  |  |  |  |  |  |
| Low -> Low | 14/912 | Reference |  | 37/442 | Reference |  |
| Low -> Medium | 18/648 | 1.64 (0.81-3.30) | 0.1686 | 20/280 | 0.80 (0.46-1.38) | 0.4215 |
| Low -> High | 6/251 | 1.51 (0.58-3.95) | 0.4037 | 4/68 | 0.64 (0.22-1.81) | 0.3963 |
| Medium -> Low | 11/415 | 1.42 (0.64-3.15) | 0.3846 | 21/175 | 1.20 (0.69-2.07) | 0.5158 |
| Medium -> Medium | 14/597 | 1.33 (0.63-2.81) | 0.4512 | 22/359 | 0.72 (0.42-1.23) | 0.226 |
| Medium -> High | 5/352 | 0.84 (0.30-2.34) | 0.7328 | 17/226 | 0.63 (0.34-1.14) | 0.1249 |
| High -> Low | 12/250 | **2.68 (1.23-5.84)** | 0.013 | 7/62 | 1.37 (0.60-3.13) | 0.45 |
| High -> Medium | 16/621 | 1.31 (0.64-2.72) | 0.4608 | 28/281 | 0.62 (0.37-1.04) | 0.0707 |
| High -> High | 14/772 | 0.97 (0.46-2.07) | 0.9442 | 23/454 | **0.44 (0.26-0.77)** | 0.0036 |

Cox proportional hazard models adjusted for age (5-year groups), sex (male or female), total energy intake, education level (high school diploma or not), household income (only in HRS), physical activity (cohort-specific categories), current smoking status (yes, no), BMI categories (underweight or normal weight, overweight, and obesity), hypertension (yes or no), diabetes (yes or no), stroke (yes or no), hypercholesterolemia (yes or no, not available in HRS), and cardiovascular diseases (yes or no).

**Supplemental Table 6.** Multivariable adjusted hazard ratios (95% confidence intervals) for incident dementia according to tertiles of Mediterranean-DASH Intervention for Neurodegenerative Delay (MIND) diet score by subgroups of participants

|  | **Whitehall II Study** | | | **Health and Retirement Study** | | | **Framingham offspring study** | | | **Pooled HR (95% CI)** |
| --- | --- | --- | --- | --- | --- | --- | --- | --- | --- | --- |
|  | **Cases/N** | **HR (95% CI)** | **P-value** | **Cases/N** | **HR (95% CI)** | **P-value** | **Cases/N** | **HR (95% CI)** | **P-value** |  |
| **Sex** |  |  |  |  |  |  |  |  |  |  |
| **Female** | 81/2582 |  |  | 192/3965 |  |  | 142/1663 |  |  |  |
| Tertile 1 | 24/664 | Reference |  | 70/1255 | Reference |  | 45/435 | Reference |  | Reference |
| Tertile 2 | 30/781 | 1.16 (0.67-2.01) | 0.5873 | 55/1096 | 0.98 (0.68-1.41) | 0.929 | 46/560 | 0.89 (0.58-1.36) | 0.5825 | 0.98 (0.76-1.25) |
| Tertile 3 | 27/1137 | 0.69 (0.39-1.23) | 0.2095 | 67/1614 | 0.87 (0.61-1.25) | 0.4489 | 51/668 | 0.76 (0.49-1.17) | 0.2055 | 0.79 (0.62-1.02) |
| per 3-unit increment | 81/2582 | 0.67 (0.43-1.07) | 0.0918 | 192/3965 | 0.80 (0.62-1.03) | 0.0809 | 142/1663 | 0.74 (0.51-1.06) | 0.1018 | 0.75 (0.62-0.91) |
| **Male** | 141/5778 |  |  | 146/2793 |  |  | 100/1382 |  |  |  |
| Tertile 1 | 46/2148 | Reference |  | 67/1089 | Reference |  | 44/562 | Reference |  | Reference |
| Tertile 2 | 45/1921 | 0.93 (0.62-1.41) | 0.7417 | 45/856 | 0.94 (0.63-1.40) | 0.7719 | 42/489 | 1.10 (0.71-1.70) | 0.6794 | 0.98 (0.77-1.24) |
| Tertile 3 | 50/1709 | 1.15 (0.76-1.74) | 0.5 | 34/848 | 0.79 (0.51-1.22) | 0.2833 | 14/331 | 0.44 (0.23-0.84) | 0.0122 | 0.77 (0.46-1.29) |
| per 3-unit increment | 141/5778 | 1.13 (0.78-1.65) | 0.518 | 146/2793 | 0.86 (0.64-1.15) | 0.3164 | 100/1382 | 0.69 (0.46-1.03) | 0.068 | 0.88 (0.68-1.13) |
| **Baseline Age** |  |  |  |  |  |  |  |  |  |  |
| **<70 years** | 75/6238 |  |  | 90/4043 |  |  | 51/2115 |  |  |  |
| Tertile 1 | 20/2185 | Reference |  | 46/1409 | Reference |  | 22/705 | Reference |  | Reference |
| Tertile 2 | 26/1995 | 1.44 (0.81-2.57) | 0.2139 | 23/1125 | 0.73 (0.43-1.21) | 0.2179 | 20/733 | 0.74 (0.40-1.38) | 0.339 | 0.91 (0.59-1.42) |
| Tertile 3 | 29/2058 | 1.40 (0.76-2.55) | 0.279 | 21/1509 | 0.61 (0.35-1.06) | 0.0787 | 9/677 | 0.37 (0.16-0.83) | 0.0162 | 0.70 (0.33-1.47) |
| per 3-unit increment | 75/6238 | 1.33 (0.80-2.20) | 0.2684 | 90/4043 | 0.64 (0.44-0.93) | 0.0186 | 51/2115 | 0.46 (0.26-0.82) | 0.0091 | 0.73 (0.40-1.32) |
| **>=70 years** | 147/2122 |  |  | 248/2715 |  |  | 191/930 |  |  |  |
| Tertile 1 | 50/627 | Reference |  | 91/935 | Reference |  | 67/292 | Reference |  | Reference |
| Tertile 2 | 49/707 | 0.83 (0.56-1.25) | 0.3756 | 77/827 | 1.03 (0.75-1.41) | 0.8485 | 68/316 | 0.89 (0.63-1.25) | 0.4968 | 0.92 (0.75-1.13) |
| Tertile 3 | 48/788 | 0.78 (0.52-1.17) | 0.2288 | 80/953 | 0.93 (0.67-1.28) | 0.6437 | 56/322 | 0.67 (0.46-0.99) | 0.0433 | 0.79 (0.64-0.98) |
| per 3-unit increment | 147/2122 | 0.77 (0.54-1.09) | 0.1437 | 248/2715 | 0.88 (0.70-1.10) | 0.2485 | 191/930 | 0.73 (0.54-0.98) | 0.038 | 0.81 (0.68-0.95) |
| **Current smoking status** |  |  |  |  |  |  |  |  |  |  |
| **Smokers** | 22/844 |  |  | 36/724 |  |  | 24/387 |  |  |  |
| Tertile 1 | 10/437 | Reference |  | 22/387 | Reference |  | 10/189 | Reference |  | Reference |
| Tertile 2 | 6/213 | 0.91 (0.31-2.72) | 0.8694 | 9/183 | 1.15 (0.51-2.59) | 0.7441 | 7/117 | 0.66 (0.22-2.03) | 0.4716 | 0.93 (0.53-1.63) |
| Tertile 3 | 6/194 | 1.53 (0.56-4.14) | 0.4062 | 5/154 | 0.81 (0.29-2.24) | 0.681 | 7/81 | 1.48 (0.43-5.14) | 0.5371 | 1.20 (0.64-2.23) |
| per 3-unit increment | 22/844 | 1.78 (0.72-4.37) | 0.2086 | 36/724 | 0.86 (0.45-1.64) | 0.6389 | 24/387 | 0.84 (0.31-2.26) | 0.7243 | 1.03 (0.65-1.65) |
| **Non-smokers** | 200/7516 |  |  | 302/6034 |  |  | 218/2658 |  |  |  |
| Tertile 1 | 60/2375 | Reference |  | 115/1957 | Reference |  | 79/808 | Reference |  | Reference |
| Tertile 2 | 69/2489 | 1.00 (0.70-1.41) | 0.98 | 91/1769 | 0.91 (0.69-1.21) | 0.5228 | 81/932 | 0.88 (0.64-1.20) | 0.4176 | 0.92 (0.77-1.10) |
| Tertile 3 | 71/2652 | 0.89 (0.62-1.27) | 0.5249 | 96/2308 | 0.82 (0.62-1.10) | 0.1808 | 58/918 | 0.58 (0.40-0.82) | 0.0025 | 0.76 (0.60-0.95) |
| per 3-unit increment | 200/7516 | 0.84 (0.62-1.14) | 0.2683 | 302/6034 | 0.80 (0.65-0.97) | 0.0269 | 218/2658 | 0.65 (0.50-0.86) | 0.0027 | 0.76 (0.65-0.87) |
| **Body weight status** |  |  |  |  |  |  |  |  |  |  |
| **Non-overweight** | 95/3305 |  |  | 104/1441 |  |  | 88/941 |  |  |  |
| Tertile 1 | 30/1110 | Reference |  | 46/477 | Reference |  | 30/259 | Reference |  | Reference |
| Tertile 2 | 31/1068 | 1.14 (0.69-1.88) | 0.6105 | 25/387 | 0.82 (0.49-1.37) | 0.4383 | 26/314 | 0.71 (0.41-1.24) | 0.2298 | 0.88 (0.65-1.19) |
| Tertile 3 | 34/1127 | 0.87 (0.51-1.47) | 0.5987 | 33/577 | 0.89 (0.54-1.47) | 0.646 | 32/368 | 0.74 (0.43-1.30) | 0.2939 | 0.83 (0.61-1.12) |
| per 3-unit increment | 95/3305 | 0.82 (0.53-1.25) | 0.3557 | 104/1441 | 0.79 (0.56-1.13) | 0.2016 | 88/941 | 0.75 (0.48-1.17) | 0.2085 | 0.78 (0.62-0.99) |
| **Overweight** | 127/5055 |  |  | 234/5317 |  |  | 154/2104 |  |  |  |
| Tertile 1 | 40/1702 | Reference |  | 91/1867 | Reference |  | 59/738 | Reference |  | Reference |
| Tertile 2 | 44/1634 | 0.93 (0.60-1.44) | 0.7498 | 75/1565 | 0.99 (0.72-1.36) | 0.9526 | 62/735 | 1.00 (0.69-1.44) | 0.9987 | 0.97 (0.79-1.20) |
| Tertile 3 | 43/1719 | 1.01 (0.65-1.56) | 0.966 | 68/1885 | 0.78 (0.56-1.09) | 0.1455 | 33/631 | 0.52 (0.33-0.83) | 0.0052 | 0.74 (0.52-1.05) |
| per 3-unit increment | 127/5055 | 1.02 (0.69-1.50) | 0.9344 | 234/5317 | 0.81 (0.64-1.01) | 0.066 | 154/2104 | 0.64 (0.46-0.90) | 0.0096 | 0.79 (0.64-0.98) |

Cox proportional hazard models adjusted for age (5-year groups), sex (male or female), total energy intake, education level (high school diploma or not), household income (only in HRS), physical activity (cohort-specific categories), current smoking status (yes, no), BMI categories (underweight or normal weight, overweight, and obesity), hypertension (yes or no), diabetes (yes or no), stroke (yes or no), hypercholesterolemia (yes or no, not available in HRS), and cardiovascular diseases (yes or no).

**Supplemental Table 7**. Multivariable adjusted hazard ratios (95% confidence intervals) for incident dementia according of Mediterranean-DASH Intervention for Neurodegenerative Delay (MIND) diet score in sensitivity analyses

|  | **Whitehall II Study** | | **Health and Retirement Study** | | **Framingham offspring study** | | **Pooled HR (95% CI)** | **P-heterogeneity** |
| --- | --- | --- | --- | --- | --- | --- | --- | --- |
|  | **Cases/N** | **HR (95% CI)** | **Cases/N** | **HR (95% CI)** | **Cases/N** | **HR (95% CI)** |  |  |
| **Alternative cutoffs** | 222/8360 |  | 338/6758 |  | 242/3045 |  |  |  |
| 0-6.9 | 46/1676 | Reference | 164/2971 | Reference | 65/674 | Reference |  |  |
| 7.0-8.9 | 107/4092 | 0.88 (0.62-1.25) | 128/2437 | 1.04 (0.82-1.33) | 112/1372 | 0.77 (0.56-1.05) | 0.91 (0.75-1.10) | 0.3192 |
| 9.0-15.0 | 69/2592 | 0.82 (0.56-1.21) | 46/1350 | 0.77 (0.55-1.09) | 65/999 | 0.55 (0.38-0.79) | 0.70 (0.55-0.89) | 0.2726 |
| **Adjusted for depressive status** | 222/8360 |  | 338/6758 |  | 237/2997 |  |  |  |
| Tertile 1 | 70/2812 | Reference | 137/2344 | Reference | 89/982 | Reference |  |  |
| Tertile 2 | 75/2702 | 1.01 (0.73-1.40) | 100/1952 | 0.97 (0.74-1.26) | 85/1037 | 0.89 (0.66-1.21) | 0.95 (0.80-1.13) | 0.8435 |
| Tertile 3 | 77/2846 | 0.96 (0.69-1.35) | 101/2462 | 0.84 (0.64-1.11) | 63/978 | 0.62 (0.44-0.87) | 0.80 (0.63-1.00) | 0.1815 |
| per 3-unit increment | 222/8360 | 0.94 (0.70-1.26) | 338/6758 | **0.82 (0.68-1.00)** | 237/2997 | 0.68 (0.52-0.89) | 0.80 (0.69-0.92) | 0.2698 |
| **Lagged for 2 years** | 220/8358 |  | 338/6758 |  | 217/3020 |  |  |  |
| Tertile 1 | 68/2810 | Reference | 137/2344 | Reference | 76/984 | Reference |  |  |
| Tertile 2 | 75/2702 | 1.03 (0.74-1.43) | 100/1952 | 0.95 (0.73-1.25) | 81/1042 | 0.95 (0.69-1.31) | 0.97 (0.81-1.15) | 0.92 |
| Tertile 3 | 77/2846 | 0.97 (0.69-1.36) | 101/2462 | 0.83 (0.63-1.09) | 60/994 | 0.67 (0.47-0.97) | 0.82 (0.68-0.98) | 0.3347 |
| per 3-unit increment | 220/8358 | 0.93 (0.69-1.24) | 338/6758 | **0.82 (0.68-0.99)** | 217/3020 | 0.76 (0.57-1.00) | 0.82 (0.72-0.95) | 0.6272 |
| **Excluding stroked participants** | 209/8179 |  | 282/6231 |  | 231/2986 |  |  |  |
| Tertile 1 | 65/2759 | Reference | 110/2117 | Reference | 83/975 | Reference |  |  |
| Tertile 2 | 72/2638 | 1.04 (0.74-1.47) | 88/1814 | 1.06 (0.79-1.42) | 85/1030 | 0.93 (0.68-1.27) | 1.00 (0.84-1.20) | 0.8192 |
| Tertile 3 | 72/2782 | 0.94 (0.67-1.34) | 84/2300 | 0.89 (0.66-1.21) | 63/981 | 0.66 (0.47-0.93) | 0.82 (0.67-1.01) | 0.2898 |
| per 3-unit increment | 209/8179 | 0.93 (0.69-1.26) | 282/6231 | 0.85 (0.69-1.04) | 231/2986 | 0.71 (0.54-0.93) | 0.82 (0.71-0.95) | 0.3959 |
| **Adjusted for continuous age** |  |  | 338/6758 |  | 242/3045 |  |  |  |
| Tertile 1 |  |  | 137/2344 | Reference | 89/997 | Reference |  |  |
| Tertile 2 |  |  | 100/1952 | 0.97 (0.74-1.27) | 88/1049 | 0.91 (0.67-1.23) |  |  |
| Tertile 3 |  |  | 101/2462 | 0.83 (0.63-1.10) | 65/999 | 0.64 (0.45-0.89) |  |  |
| per 3-unit increment |  |  |  | 0.81 (0.67-0.98) |  | 0.67 (0.51-0.88) |  |  |

Cox proportional hazard models were adjusted for age (5-year groups), sex (male or female), total energy intake, education level (high school diploma or not), household income (only in HRS), physical activity (cohort-specific categories), current smoking status (yes, no), BMI categories (underweight or normal weight, overweight, and obesity), hypertension (yes or no), diabetes (yes or no), stroke (yes or no), hypercholesterolemia (yes or no, not available in HRS), and cardiovascular diseases (yes or no), unless specified.

**Supplemental Table 8**. Characteristics of included studies in the meta-analysis

| **Author, year** | **Study Population** | **Country** | **Follow-up (y)** | **Sample size** | **Age Range (years)** | **Assessment of Diet** | **Ascertainment of Cases** | **No of cases** | **Categories of Exposure** | **Relative Risks (95% CI)** | **Covariates in Multivariable Model** |
| --- | --- | --- | --- | --- | --- | --- | --- | --- | --- | --- | --- |
| Morris, 2015 | MAP | US | 4.5 | 789 | 58-98 | FFQ | Clinical diagnosis | 135 | Tertiles | 0.47, (0.29, 0.76) for the highest tertile | Age, sex, education, APOE ε4 (any), participation in cognitively stimulating activities, physical activity, and total energy intake. |
| de Crom, 2022 | RS | Netherlands | 15.6 (B I, 5.9 (B II) | 5375 (B I), 2861 (B II), largely non-overlap | ≥55 | FFQ | Clinical diagnosis | 1188 (B I) and 248 (B II) | Per SD | 0.99 (0.94–1.05) for baseline I 0.79 (0.70–0.91) for baseline II | Sex, age, age^2, educational attainment, smoking status, physical activity, daily energy intake, body mass index, diabetes, hypercholesterolemia, and hypertension |
| Zhang, 2022 | UK Biobank | UK | 10.5 | 187,783 | ≥40 | 24h diet recall | HES linkage | 1,363 | Tertiles | 0.87 (0.76-0.99) for the highest tertile | Age, sex, education level, Townsend deprivation index, body mass index, smoking status, alcohol consumption, regular physical activity, time on watching TV, sleep duration, family history of AD, APOE genotype, and cardiovascular disease, cancer, and diabetes at baseline |
| Vu, 2022 | MAP, CHAP, WHIMS | US | Not reported | 2449 (CHAP), 725 (MAP), 5308 (WHIMS) | ≥65 | FFQ | Clinical diagnosis | 222 (MAP), 951 (WHIMS), 67 (CHAP-White), and 109 (CHAP-Black) | Tertiles | 0.63 (0.42, 0.92), 0.80 (0.72, 0.89), 1.23 (0.47, 3.18), 1.48 (0.51, 4.27) for the highest tertiles in MAP, WHIMS, CHAP-White and CHAP-Black, respectively | Age, sex, education, income, global cognition score, late-life cognitive activity, history of diabetes, hypertension, stroke and heart disease, smoking, calorie intake, BMI, depressive symptoms and physical activity. |
| Thomas, 2022 | 3C-Bordeaux | France | 9.7 | 1412 | ≥65 | FFQ | Clinical diagnosis | 356 | Tertiles | 0.73 (0.55–0.97) for the highest tertile | Age, body mass index (BMI), total energy intake, depressive symptoms, sex, educational level, tobacco consumption and engagement in regular physical activity, APOE ε4 genotype, history of cardiovascular or cerebrovascular disease, hypertension, hypercholesterolemia, and diabetes |

**Supplemental Table 9**. Risk of bias of the included studies in the Newcastle-Ottawa scale

|  | **Selection** | | | |  | **Comparability** | |  | **Outcome** | | |  | **Total** |
| --- | --- | --- | --- | --- | --- | --- | --- | --- | --- | --- | --- | --- | --- |
|  | **Representativeness of the exposed cohort** | **Selection of the non-exposed cohort** | **Ascertainment of exposure** | **Outcome of interest not present at start of the study** |  | **Control for primary confounders** | **Control for secondary confounders** |  | **Assessment of outcome** | **Duration of follow-up** | **Adequacy of follow-up** |  |  |
| Morris, 2015 | 1 | 1 | 1 | 1 |  | 1 | 1 |  | 1 | 0 | 1 |  | 8 |
| de Crom, 2022 | 1 | 1 | 1 | 1 |  | 1 | 1 |  | 1 | 1 | 1 |  | 9 |
| Zhang, 2022 | 0 | 1 | 1 | 1 |  | 1 | 0 |  | 1 | 1 | 1 |  | 7 |
| Vu, 2022 | 1 | 1 | 1 | 1 |  | 1 | 1 |  | 1 | 0 | 1 |  | 8 |
| Thomas, 2022 | 1 | 1 | 1 | 1 |  | 1 | 1 |  | 1 | 0 | 1 |  | 9 |

Representativeness of the exposed cohort: 1 if community-based, 0 if volunteer-based.

Selection of the non-exposed cohort: 1 if the non-exposed cohort is selected from the same population as the exposed group.

Ascertainment of exposure: 1 if repeated measurements are conducted, 0 if not.

Outcome of interest not present at start of the study: 1 if excluding prevalent dementia at baseline, 0 if not.

Control for primary confounders: 1 if controlled for age, sex, socioeconomic status, lifestyle factors, and other dietary confounders (such as total energy intake); 0 if missing any.

Assessment of outcome: 1 if diagnosed by doctors or with validated algorithm, 0 if not.

Duration of follow-up: 1 if >=10 years, 0 if <10 years or not reported.

Adequacy of follow-up: 1 if loss to follow-up <30%, 0 if not.

**Supplemental Figure 1**. Funnel plot in the meta-analysis

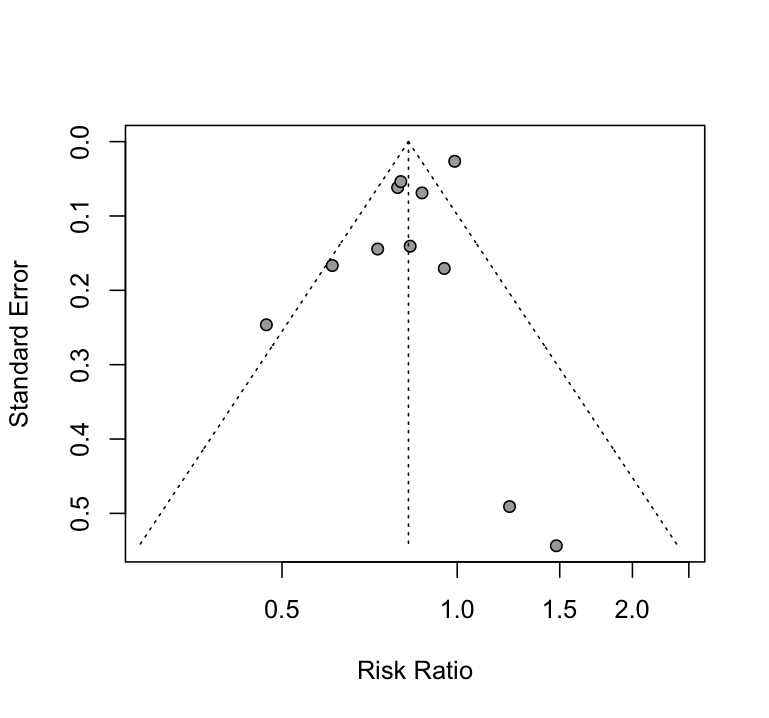
